# Overcrowded Housing Reduces COVID-19 Mitigation Measures and Lowers Emotional Health Among San Diego Refugees from September to November of 2020

**DOI:** 10.1101/2023.01.20.23284851

**Authors:** Ashkan Hassani, Vinton Omaleki, Jeanine Erikat, Elizabeth Frost, Samantha Streuli, Ramla Sahid, Homayra Yusufi, Rebecca Fielding-Miller

## Abstract

Refugee communities are vulnerable to housing insecurity, which drives numerous health disparity outcomes in a historically marginalized population. The COVID-19 pandemic has only worsened the ongoing affordable housing crisis in the United States while continuing to highlight disparities in health outcomes across populations. We conducted interviewer-administered surveys with refugee and asylum seekers in San Diego County at the height of the COVID-19 pandemic to understand the social effects and drivers of COVID-19 in one of the largest refugee communities in the United States. Staff from a community-based refugee advocacy and research organization administered the surveys from September - November 2020. 544 respondents participated in the survey, which captured the diversity of the San Diego refugee community including East African (38%), Middle Eastern (35%), Afghan (17%), and Southeast Asian (11%) participants. Nearly two-thirds of respondents (65%) reported living in overcrowded conditions (> <u>1 individual per room</u>) and 30% in severely crowded conditions (≥ 1.5 individuals per room). Respondents living in affordable housing units or receiving section 8 housing vouchers had a 66% lower probability of living in severely crowded settings (aOR:0.34, 95% CI:0.19– 0.61). Refugees living in overcrowded and severely overcrowded housing had more than twice the odds to have not accessed COVID-19 testing since the pandemic began (OR: 2.28, 95% CI: 1.38 - 3.78) and had nearly 4 times the odds to report lower emotional health (OR: 3.90, 95% CI: 2.62 - 5.82). Longer United States residency was associated with a 7% reduction in the odds of living in crowded housing per additional year (aOR:0.93, 95% CI:0.90–0.97). Overcrowding housing is a structural burden that reduces COVID-19 risk mitigation behaviors. Improved access to affordable housing units or receiving vouchers could reduce overcrowded housing in vulnerable refugee communities.

## Introduction

### Background

The immigrant and refugee community in the United States has experienced high rates of morbidity and mortality as a result of the COVID-19 pandemic (1,2). Globally, in high-income countries, foreign-born residents are at higher risk of COVID-19 transmission, have lower access to testing, and have higher rates of hospitalization and mortality due to COVID-19 compared to native-born residents (2). In San Diego County, one analysis of death certificates found that although foreign-born residents comprise 23% of county residents, they accounted for 40% of all deaths attributed to COVID-19 between March 22, 2020 and March 22, 2021 (1). Increased COVID-19 vulnerability in the immigrant and refugee community stems from a number of social and structural factors: Recent immigrants are more likely to work low-wage, frontline jobs, which carry a higher risk of disease acquisition and transmission, and are less likely to provide paid sick leave (3,4). Concerns about the public charge and a lack of documentation on immigration status may also deter individuals from seeking timely care when an infection is suspected (5,6). As a consequence of working low-wage jobs and racially discriminatory housing practices, immigrants are 4 times more likely to live in crowded housing (7). This can exacerbate secondary attack rates within household units and present challenges for safe isolation (7).

Overcrowding in homes - sometimes referred to as ‘hidden homelessness’ has been highlighted by activists and researchers as a major issue in the refugee community in the United States (8–10). Data from Canada and Europe document the increased risk of infectious disease among refugee communities living in overcrowded housing (8,11). Access to affordable housing (defined by the Department of Housing and Urban Development as ‘housing that costs no more than 30% of your income) is associated with benefits such as improved food security, health care access, reduced stress, housing stability, improved indoor environmental health, and increased mental health (12). Despite these benefits, access to affordable housing is severely limited by a backlogged federal Section 8 voucher program for affordable housing and state and local policies that have restricted new constructions of affordable housing and banned public housing (13).

### Objectives

Immigrant and refugee communities in the United States have been raising the issue of housing as a health and human rights issue for many years (10,14). As part of their biennial community survey conducted during the first peak of the pandemic on the state of the refugee community in San Diego County, the Partnership for the Advancement of New Americans (PANA) partnered with the University of California, San Diego (UCSD) to better understand the link between housing and COVID-19 vulnerability among refugee communities in Southern California.

## Methods

### Study design and setting

The Partnership for the Advancement of New Americans (PANA) is a research, public policy, and community organizing hub that serves refugees and asylum seekers in San Diego County (10). Data for the present cross-sectional study were collected between September 2020 and November 2020 as part of PANA’s biannual community survey. The survey was designed in English and translated into Arabic and Spanish by bilingual study staff. Interviewers were provided with a list of PANA members and the survey was subsequently administered over the phone and in person by trained research assistants in Arabic, Burmese, Dari, English, Karenni, Oromo, Pashto, Somali, Spanish, and Swahili. Up to two household members could be interviewed. However, interviewers tracked whether an individual had completed a survey or not. Data were collected as part of a programmatic report intended to provide a general overview of the community. As such, we did not conduct *a priori* power calculations focused on one specific outcome.

Interviewers entered participant answers using Qualtrics software (Qualtrics ver. 2021). The survey contained 83 questions regarding demographics, COVID-19, housing, employment, health, children, belonging, and resilience. The survey took approximately 30 minutes to complete and up to 1 hour if it was being translated. The study team met weekly to discuss any issues with translation and address questions raised by survey administrators. Consistent with our participatory action approach, all data collection and analyses were conducted in conversation with PANA staff and guided by their lived expertise and organizational policy priorities. Variables for the present analyses were chosen in consultation with PANA leadership to address the primary research question: What is the association between housing and COVID-19 vulnerability? Vulnerability was broadly understood as both healthcare access and the broader mental health impact of the epidemic (15). The full report with all variables can be found elsewhere (10).

### Participants

Participants were recruited using a convenience sampling approach. Individuals were eligible to participate in the study if they were over the age of 18, able to participate in a language spoken by a trained research assistant, willing to provide informed consent, and part of the refugee community. “Refugee community member” was construed broadly and included individuals who had arrived as refugees, asylum seekers, or their American-born children. PANA staff contacted all individuals for whom the organization had provided services or with whom they had engaged in community organizing efforts in the previous 5 years. Each staff interviewer was assigned a list of people based on language. Research assistants also recruited participants in person in shopping areas that members of the refugee community typically frequent. Staff interviewed and conducted the survey by phone with each individual from the convenience sample - ensuring no duplication. Interviews were limited to no more than two individuals per household (typically a parent and an adult child). All participants who completed the survey were given a $20 gift card to thank them for their time and expertise.

### Measures and analyses

Within the survey, participants were asked about the total number of individuals living in their homes and the total number of rooms in the home. In accordance with the California Department of Public Health (CDPH), overcrowding was defined as more than one individual per room. Severe overcrowding was defined as 1.5 individuals per room (16). For example, a traditional 2-bedroom apartment would have 4 distinct rooms, including the kitchen and living room, and it would require 6 inhabitants to be classified as “severe overcrowding.” Other explanatory variables included access to affordable housing (Are you in an affordable housing unit, or receiving a section 8 voucher to help with the rent?), access to a diagnostic test for COVID-19 (Have you ever gotten a test for COVID-19?), and self-rated emotional health with 5 potential responses ranging from “never” to “always” (How often have you been bothered by emotional problems such as feeling anxious, depressed or irritable?). In consultation with PANA leadership, the study team decided to dichotomize the emotional health variable for analysis and compare participants who reported ‘never’ experiencing emotional problems vs all others. Demographic variables such as income from the previous week (Have you worked for money in the previous week), age, year of arrival to the United States, gender (male or female options only), cohabiting with a partner, number of children (How many children under the age of 18 are you responsible for?), and family size.

We first examined basic univariate frequencies as a study team to develop a basic understanding of sample demographics and the prevalence of indicators of interest within the sample. We then used chi-square and student’s t-tests to test the hypothesis that our primary predictor and covariates of interest were significantly associated with living in severely overcrowded housing. We built simple logistic regression models to measure the unadjusted odds of association with overcrowded housing and finally constructed a full multivariate regression with all variables of interest to measure the adjusted odds of association. While the emotional health item was collected using a Likert scale, it was modeled as a continuous variable as this was the most conceptually consistent way to consider an individual’s emotional well-being. We conducted a simple logistic regression to examine the associations for overcrowded housing. All analyses were conducted using Stata 16 (17).

### Ethical considerations and IRB

Informed Consent was verbally obtained from all participants by trained PANA staff before administering the Qualtrics survey. Data gathered from the questionnaires was confidential. This study was reviewed and approved by the University of California, San Diego Institutional Review Board (UCSD IRB) under project #201601SX. In consideration of the vulnerable status of our study population, we adopted a participatory action approach to this community-led project and our team from UCSD only provided technical assistance (18).

## Results

PANA staff contacted 680 community members and 544 agreed to participate in the survey, for a response rate of 80%. The mean time living within the United States was 12 years, and gender was approximately evenly distributed (Table 1). Just over 1 in 4 participants reported that they had engaged in work for money in the previous two weeks. Living in crowded or severely crowded housing conditions was common: participants reported an average of 1.5 individuals per room, with 29.9% (n=160) living in severely overcrowded conditions. Thirty-two percent (n=172) lived in affordable housing units or utilized section 8 vouchers. At the time the study was conducted (September - November 2020), 23% of participants had ever accessed a diagnostic test for COVID-19 (n=123). Approximately 37% of participants reported that they sometimes, half the time, most of the time, or always experienced emotional problems like anxiety or depression.

**Table 1.** Sample Demographics of the refugee participants in San Diego, CA, sampled between November 2020-December 2020. Includes age, average years in the United States, English spoken at home, and gender stratified by region of origin.

| Region | East Africa | Syria | Afghanistan | Southeast Asia | Total |
| --- | --- | --- | --- | --- | --- |
|  | n=205 | n=184 | n=90 | n=56 | N=535 |
| **Age** |  |  |  |  |  |
| Mean (SD) | 38.5 (16.5) | 42.5 (10.1) | 35.8 (10.3) | 37.0 (14.8) | 39.3 (13.7) |
| Range | 15.0 - 96.0 | 15.0 - 68.0 | 14.0 - 70.0 | 14.0 - 68.0 | 14.0 - 96.0 |
| **Years in the United States** |  |  |  |  |  |
| Median | 17.0 | 4.0 | 4.0 | 7.0 | 5.0 |
| Range | 1.0 - 35.0 | 1.0 - 43.0 | 0.0 - 35.0 | 1.0 - 26.0 | 0.0 - 43.0 |
| **English spoken at home** | 82 (40.0%) | 17 (9.2%) | 73 (81.1%) | 14 (25.0%) | 186 (34.8%) |
| **Female** | 150 (74.6%) | 38 (20.7%) | 11 (12.2%) | 41 (73.2%) | 240 (45.2%) |

In simple logistic regressions, living in severely overcrowded housing was significantly associated with having never accessed a test for COVID-19 (OR: 2.28, 95% CI: 1.38 - 3.78) and higher odds of reporting lower emotional health (OR: 3.90, 95% CI: 2.62 - 5.82). Individuals who reported accessing affordable housing were 81% lower odds of reporting living in severely crowded conditions (OR: 0.19, 95% CI: 0.12 - 0.33). Older age and more years living in the United States also appeared to be protective against overcrowding (Table 2).

**Table 2.** Overcrowded vs. Severely Overcrowded Housing among participants in San Diego, CA from September to November 2020.

|  | Total<br>(n=535) | Not severely<br>overcrowded<br>(n = 375) | Severely<br>overcrowded<br>(n = 160) |  |
| --- | --- | --- | --- | --- |
| *Measures* | *Mean (SD)* | *Mean (SD)* | *Mean (SD)* | *p-value* |
| People per room | 1.5 (0.9) | 1.1 (0.3) | 2.5 (1.0) | <0.001 |
| Years in the United States | 10.1 (8.5) | 12.0 (8.8) | 6.0 (6.1) | <0.001 |
| Age | 39.0 (13.8) | 39.8 (14.7) | 37.1 (11.2) | 0.040 |
|  | *n (%)* | *n (%)* | *n (%)* |  |
| Never accessed COVID-19 test | 410 (77.1) | 273 (73.2) | 137 (86.2) | 0.001 |
| How often are you bothered by emotional problems like anxiety or depression? |  |  |  | 0.001 |
| Always | 21 (3.9) | 14 (3.7) | 7 (4.4) |  |
| Most of the time | 19 (3.6) | 13 (3.5) | 6 (3.8) |  |
| About half the time | 17 (3.2) | 15 (4.0) | 2 (1.3) |  |
| Sometimes | 195 (36.7) | 99 (26.5) | 96 (60.8) |  |
| Never | 280 (52.6) | 233 (62.3) | 47 (29.7) |  |
| Affordable housing | 172 (32.2) | 153 (40.9) | 19 (11.9) | <0.001 |
| Employed | 144 (27.2) | 95 (25.6) | 49 (31.0) | 0.200 |
| Female | 238 (44.9) | 189 (50.9) | 49 (30.8) | <0.001 |

The fully adjusted model is shown in Table 3. The model had a relatively good fit, with a pseudo-r-squared value of 0.20, no specification errors identified using the *link test* command in Stata (_hatsq p = 0.68), a non-significant Hosmer-Lemeshow goodness-of-fit test (p=0.78)., Individuals who had never accessed a test for COVID-19 had 2.15 times the adjusted odds of living in severely crowded conditions (95% CI: 1.19 - 3.88). Individuals who reported lower emotional health had 4 times greater odds of living in severely overcrowded housing (aOR: 4.37, 95% CI: 2.72 - 7.03). Access to affordable housing decreased the odds of living in severely crowded housing by over 60% (95% CI: 0.21 - 0.75). Older age and longer time spent in the United States were also associated with decreased odds of living in severely crowded housing conditions.

**Table 3.** Simple and adjusted odds of living in severely overcrowded housing vs. not living in severely overcrowded housing

|  | Odds Ratio | 95% CI | Adjusted Odds Ratio | 95% CI |
| --- | --- | --- | --- | --- |
| Never accessed COVID-19 test* | 2.28 | 1.38 - 3.78 | 2.15 | 1.19 - 3.88 |
| Lower emotional health* | 3.90 | 2.62 - 5.82 | 4.37 | 2.72 - 7.03 |
| Affordable housing* | 0.19 | 0.12 - 0.33 | 0.39 | 0.21 - 0.75 |
| Employed | 1.31 | 0.87 - 1.97 | 0.97 | 0.59 - 1.60 |
| Family size* | 1.32 | 1.20 - 1.44 | 1.30 | 1.15 - 1.47 |
| Years in the United States* | 0.89 | 0.86 - 0.92 | 0.94 | 0.91 - 0.98 |
| Age | 0.99 | 0.98 - 1.0 | 0.97 | 0.95 - 0.99 |
| Female* | 0.43 | 0.29 - 0.64 | 0.83 | 0.49 - 1.01 |

## Discussion

### Key Results

We found that within the refugee community, living in severely overcrowded housing was significantly associated with a decreased probability of accessing a COVID-19 test and an increased probability of reporting lower emotional health. Individuals who had never accessed a test for COVID-19 were more likely to live in severely crowded housing and significantly more likely to report lower emotional health.

Meanwhile, those who reported accessing affordable housing were significantly less likely to report living in severely crowded conditions. Older age and more years lived in the United States were also associated with decreased odds of severely crowded housing conditions.

### Interpretation

The role of housing during the COVID-19 pandemic has focused mainly on how having shelter can mitigate risks of SAR-CoV-2 transmission and the need for moratoriums on evictions (19,20). Increased risk of exposure to COVID-19 is already widely associated with overcrowded housing (21–27). However, less well documented are the ways in which living in overcrowded housing may be linked to decreased ability for risk mitigation behaviors, such as accessing COVID-19 diagnostic testing. If a person is living in severely overcrowded housing without the ability or resources to properly quarantine or isolate then the results of a COVID-19 test may not have much impact on behavior, which can diminish incentives for testing. Furthermore, the quality of housing and the density of inhabitants per domicile during the pandemic has garnered less attention, especially when regarding public health policy (18).

There is a significant association in the link between lower emotional/mental health and crowded housing among the population we studied. Discussing mental health is highly stigmatized in this community, and the study team agreed that disclosing any concerns related to emotional or mental health in this context was likely indicative of significant levels of emotional distress. There is evidence that overcrowded housing can have a negative impact on an individual’s mental health (28,29). Our findings add to this literature but our narrowed focus on the refugee community highlights that housing conditions can add an extra emotional and mental burden to a population already at high risk of lower emotional/mental health outcomes (30,31). This combination of the mental strains that refugees in overcrowded housing experience may require service providers to tailor their interventions.

Affordable housing significantly reduces the odds of living in severely crowded conditions. Older age and more years living in the United States also appear to be protective. However, age and years in the United States can be an effect-modifier for affordable housing. There is an average wait of 8-10 years for applicants to receive a federal section 8 housing voucher to move into an affordable home which is consistent with our data that shows newly arrived refugees are at even higher odds of overcrowding, and that the longer they live in the United States, the more likely they are to live in affordable housing (32). Our findings also suggest that refugees living in San Diego are experiencing economic factors that force multiple refugee families to share spaces meant for only one family. This is a form of ‘hidden homelessness (11). Just over 1 in 4 individuals reported engaging in work for money in the previous 2 weeks. While some of this number may be an artifact of sampling bias (i.e., those who responded were more likely to be free during the day to participate in the survey) and/or represent individuals who are full-time students or homemakers, previous work conducted by our team demonstrated that the refugee community experienced high rates of job loss during the pandemic (32). These overcrowded housing arrangements may be necessary for economic survival, but in the context of the COVID-19 pandemic, they become hot spots of risk, increasing the likelihood of acquiring, transmitting, and reinfection with the virus (33).

### Limitations

This is a cross-sectional study with a convenience sample. As such we cannot make claims about the directionality of the associations we identified, nor should our data be interpreted as representing a true prevalence. However, a full census of the refugee population in the region - or country - does not exist, making random sampling impractical if not impossible. Additionally, gender was limited to two options (male or female) to preserve statistical significance and we are aware that this may not be a true representation of the diverse population. Survey data was self-reported so it is possible certain topics were biased towards more socially acceptable answers. Furthermore, although the survey was limited to two participants per household, the survey did not record how many participants belonged in the same household. Therefore, we were unable to adjust household size as a fixed effect. However, PANA interviewers reported that in practice it was unusual for two respondents from the same household to participate in the survey.

## Conclusions

Refugees living in overcrowded and severely overcrowded housing had lower odds of accessing a COVID-19 test, which highlights a structural burden to risk mitigation behaviors in refugee populations who may be experiencing hidden homelessness. These housing conditions may also be impacting refugee emotional health. Promptly increasing access to affordable housing for vulnerable refugee populations may protect against the increased odds of COVID-19 and lower emotional health associated with overcrowding. We recommend that local governments utilize federal resettlement funding in collaboration with local refugee groups to develop and implement affordable housing plans for refugees upon arrival. This will ensure that recently resettled refugees are able to build long-term stability while improving population health in the United State’s most marginalized communities. In addition, we recommend the federal government increase the number of Section 8 vouchers available so that wait times are reduced (34). Finally, we recommend that the State of California create legislation and provide funding for permanent social housing that refugee populations can access (10). Future directions in research should look at the impact of master leasing, housing vouchers, and universal basic income on refugee housing security and general health outcomes.

## Data Availability

We are unable to directly share the data set as it is legally owned by a third-party organization - Partnership for the Advancement of New Americans (PANA). Our analyses were conducted with the permission of PANA leadership. Individuals interested in accessing the data may contact PANA with inquiries at. Please include "PANA_2020_HML_Survey" in the subject line, your affiliation, and the reason you are requesting the data set.

